# Listener effort quantifies clinically meaningful progression of dysarthria in people living with amyotrophic lateral sclerosis

**DOI:** 10.1101/2024.05.31.24308140

**Authors:** Indu Navar Bingham, Raquel Norel, Esteban G. Roitberg, Julián Peller, Marcos A Trevisan, Carla Agurto, Diego E. Shalom, Felipe Aguirre, Iair Embon, Alan Taitz, Donna Harris, Amy Wright, Katie Seaver, Stacey Sullivan, Jordan R. Green, Lyle W. Ostrow, Ernest Fraenkel, James D. Berry

## Abstract

Amyotrophic lateral sclerosis (ALS) is a neurodegenerative motor neuron disease that causes progressive muscle weakness. Progressive bulbar dysfunction causes dysarthria and thus social isolation, reducing quality of life. The Everything ALS Speech Study obtained longitudinal clinical information and speech recordings from 292 participants. In a subset of 120 participants, we measured speaking rate (SR) and listener effort (LE), a measure of dysarthria severity rated by speech pathologists from recordings. LE intra- and inter-rater reliability was very high (ICC 0.88 to 0.92). LE correlated with other measures of dysarthria at baseline. LE changed over time in participants with ALS (slope 0.77 pts/month; p<0.001) but not controls (slope 0.005 pts/month; p=0.807). The slope of LE progression was similar in all participants with ALS who had bulbar dysfunction at baseline, regardless of ALS site of onset. LE could be a remotely collected clinically meaningful clinical outcome assessment for ALS clinical trials.

## Introduction

ALS causes progressive weakness of muscles under voluntary control throughout the body. It begins in the bulbar region in approximately 25-30% of people (1). Progressive bulbar dysfunction and dysarthria eventually develops in 80-95% of people living with ALS (2). The ALS Functional Rating Scale – Revised (ALSFRS-R) is traditionally used to quantify disease progression (3), and the ALSFRS-R Self Entry (i.e., self-reported; ALSFRS-RSE) correlates highly with the ALSFRS-R and progress at a similar rate (4, 5). Thus, ALSFRS-RSE is frequently used in remote ALS studies. Despite its frequent use, the ALSFRS-R (and ALSFRS-RSE) is a blunt instrument for testing dysarthria, since only one question (Q1) asks about speech intelligibility, which is rated 0-4, and only three questions (Q1-3) focus on bulbar function (ALSFRS-R bulbar subdomain).

Quantitative motor speech (QMS) analysis describes standard analyses aimed at quantifying characteristics or features of speech such as rate, pause or articulation. QMS is conducive to remote studies because it can be implemented using speech recordings obtained in the home environment on personal electronic devices (smartphones, tablets, or computers), allowing frequent, simple data collection. In ALS, QMS has focused on speaking rate (SR; words/minute), articulation rate (AR; syllables/second), and speech pause analysis (SPA) to identify speech abnormalities, which in some instances, can be detected even before people with ALS (PALS) or their speech and language pathologists (SLPs) are aware of them (4, 6–10). However, each of these features may be insufficient to quantify ALS progression over time. Dysarthria is a complex process. It can occur due to dysfunction in any one or more of the speech subsystems (articulatory, resonatory, phonatory, respiratory) (11, 12). Furthermore, in complex degenerative disorders like ALS, each of the speech subsystems can decline at different rates, and numerous compensatory mechanisms can further complicate patterns of dysarthria. While the net effect is a decline in the overall ability to communicate, this complexity renders both traditional blunt instruments like the ALSFRS-R and specific measures of individual speech subsystem performance limited in their ability to identify and quantify clinically meaningful progression of dysarthria (8). Overall quantification of dysarthria severity in ALS may require more complex analysis and models that incorporate features from different speech subsystems simultaneously (11, 13).

SLP ratings of percent intelligibility (14), for example, have demonstrated modest reliability at quantifying the severity of dysarthria in ALS (15) and correlate with PALS self-reports of dysarthria and resulting distress (16) (17). These measures have even been used to estimate the time to loss of intelligibility in people living with ALS – approximately 32 months for people with bulbar onset ALS (14). Listener effort (LE) is a perceptual rating of the amount of work necessary for a listener to understand a speaker with disordered speech (18). It has been used extensively as a patient-reported outcome measure in studies of the impact of the clinically meaningful impact of hearing impairment (19, 20). In disorders causing dysarthria, it is an assessment made by a healthy listener to quantify the negative impact of dysarthria (18); and, it has proven to be one of the most robust overall measures of dysarthria (15).

We conducted a fully remote observational study characterizing speech in ALS. We analyzed these recordings to characterize SR and AR. Speech and language pathologists (SLPs; (DH, AW, KS, SS)) with ALS expertise then rated LE to evaluate its performance as a digital COA in ALS. Finally, we developed a machine-learning model to predict LE, the Listener Effort Prediction Model (LEPM).

Our analysis recapitulates prior results demonstrating that SR and AR can quantify aspects of dysarthria in ALS. More importantly, we demonstrate that LE is a quantifiable ALS clinical outcome assessment (COA) that correlates with ALSFRS-RSE Q1 and bulbar subdomain and SR. Furthermore, LE is more sensitive to change than these features, suggesting that it may be both a clinically meaningful and statistically powerful clinical outcome assessment in ALS trials. Finally, we show a high predictive value of the LEPM, showing that the LEPM algorithm may have promise as a digital clinical outcome assessment of dysarthria in ALS. The repository of recorded speech and de-identified clinical data from this study is now available to ALS researchers to advance speech research in ALS (https://www.everythingals.org/available-data).

## Results

### Study Enrollment and Demographics

In total, 525 participants entered the study and recorded at least 1 session, 457 recorded at least 2, 412 recorded 3 or more (range of recording numbers: 1-144). After matching all participants’ recording and clinical data and passing data through quality control, the dataset included 292 participants (136 with ALS and 156 controls) with a total of 6272 speech recording sessions for a total of 56,462 individual speech recording tasks (Figure 1).

**Figure 1:**
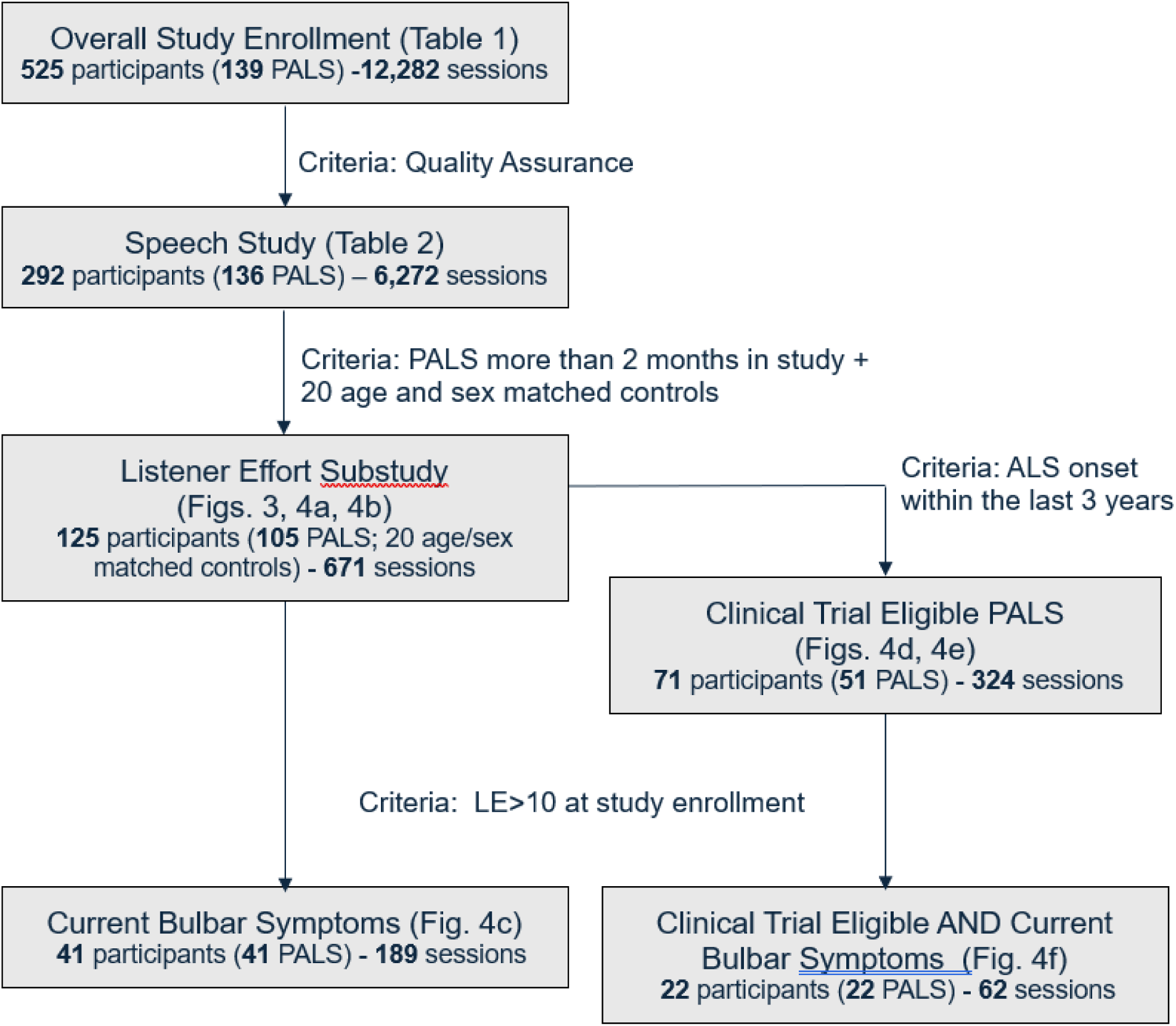
Cohort Diagram. Flow of participants from enrollment in the overall study, through quality assurance, inclusion in the overall speech study, inclusion in the Listener Effort Substudy and creation of cohorts within the LE Substudy. Criteria for inclusion in sub-studies and cohorts are noted. Cohorts described in specific Tables and Figures throughout the study are noted.

Of the 292 participants, 40.8% male and 59.2% female, and the median age was 65 (range 30-85). In this study, 91% of PALS were white, far more than reported in the US National ALS Registry of 71% (21) but similar to other ALS clinical studies (22, 23). (Table 1). The participants were enrolled from 15 states, though 27% did not provide geographic information (Supplementary Figure 1).

**Table 1:** Demographics and ALS Disease Characteristics of speech study and LE Substudy.

|  | Overall Speech Study |  |  | LE Substudy |  |  |
| --- | --- | --- | --- | --- | --- | --- |
|  | All | PALS | Ctrl | All | PALS | Ctrl |
| **N** | 292 | 136 | 156 | 125 | 105 | 20 |
| **Age (median, range)** | 65 (30-85) | 66 (32-83) | 63 (30-85) | 67 (31-85) | 67 (39-83) | 67 (31-85) |
| **Sex (n)** |  |  |  |  |  |  |
| **Female** | 173 | 67 | 106 | 60 | 50 | 10 |
| **Male** | 119 | 69 | 50 | 65 | 55 | 10 |
| **Ethnicity (n)** |  |  |  |  |  |  |
| **Non-Hispanic** | 273 | 128 | 145 | 116 | 98 | 18 |
| **Hispanic** | 14 | 5 | 9 | 6 | 5 | 1 |
| **Unknown** | 5 | 3 | 2 | 3 | 2 | 1 |
| **Race (n)** |  |  |  |  |  |  |
| **White** | 265 | 124 | 141 | 112 | 94 | 18 |
| **Asian** | 13 | 5 | 8 | 5 | 5 | 0 |
| **American Indian or Alaska Native** | 6 | 3 | 3 | 4 | 3 | 1 |
| **Black or African American** | 4 | 2 | 2 | 1 | 1 | 0 |
| **No data** | 4 | 2 | 2 | 3 | 2 | 1 |
| **Baseline ALSFRS-R (mean, std)** | 42.3 (7.7) | 36.1 (7.2) | 47.8 (0.9) | 38.1 (7.7) | 36.2 (7.0) | 47.9 (0.3) |
| **Diagnostic Delay (time between onset of weakness and diagnosis (yrs))** |  | 1.4 (1.4) |  |  | 1.4 (1.5) |  |
| **Time Since ALS Symptom Onset at Baseline (yrs)** |  | 3.9 (3.7) |  |  | 4.1 (3.8) |  |
| **Reported ALS Inheritance (n)** |  |  |  |  |  |  |
| **Familial** |  | 48 |  |  | 40 |  |
| **Sporadic** |  | 88 |  |  | 65 |  |
| **Onset location (n)** |  |  |  |  |  |  |
| **Bulbar** |  | 34 |  |  | 23 |  |
| **Non-bulbar** |  | 101 |  |  | 81 |  |
| **Unknown** |  | 1 |  |  | 1 |  |

#### Motor Speech Characteristics at Baseline and Longitudinally

As noted, quantitative motor speech analyses of speaking rate (words/minute; SR) and articulation rate (syllables/second; AR) have been used extensively in ALS speech studies. In this cohort, the average baseline SR for PALS was 145.9 words/min (SE 3.8) and for controls was 186.1 words/min (SE 1.6; p<0.001), and the average baseline AR for PALS was 3.339 syllables/sec (SE 0.085) and for controls was 4.269 syllables/sec (SE 0.036; p<0.001). Longitudinal analysis of AR for PALS shows a slope of −0.0035 units/month (SE 0.0013; p=0.010) and for controls 0.0125 units/month (SE 0.0010; p=0.23). Longitudinal analysis of SR for PALS shows a slope of −0.203 units/month (SE 0.079; p=0.011) and for controls shows a slope of 0.34 units/month (SE 0.45; p=0.45) (Supplementary Figure 2, panels a and d).

People with bulbar onset ALS had slower speech at baseline than non-bulbar onset (bulbar-onset 117.6 words/min (SE 10.6); non-bulbar 170.7 (SE 5.1); p < 0.001) (Supplementary Figure 2, panels b and e). However, those experiencing bulbar symptoms at study entry had similar speaking rate at baseline and progression of slowing over time regardless of site of ALS onset (non-bulbar with bulbar symptoms at study entry 117.6 words/min (SE 10.3), bulbar-onset 110.8 (SE 7.6), p = 0.59) (Supplementary Figure 2, panels c and f).

### LE Sub-Study

Because AR, SR and other quantitative motor speech features only characterize individual facets of dysarthria, and ALS is characterized by complex degeneration of multiple speech subsystems, we sought to identify a more robust overall measure of progressive dysarthria in ALS. While we favored LE as an overall measure of dysarthria based on our prior experience, we conducted two pilot projects to select the most robust metric for further study.

We investigated recordings of three tasks: reading the bamboo passage (24), freely describing a line drawing picture (25), and reading sentences (26). Each task was rated with 11 outcome measures (LE, Overall Dysarthria Severity, Slow Speaking Rate, Voice Strain, Consistency, Reduced Intelligibility, Articulatory Imprecision, Dysphonia Severity, Hypernasality, Reduced Breath Support, Reduced Prosody). Based on its performance in these pilot studies and our prior experience, the task selected for further study was sentence reading and the outcome measure selected was LE.

LE is denoted as a 0-100 score, with higher numbers representing more effort, thus worse dysarthria. Because LE can be derived from live interviews or speech recordings, it is both useful and practical for a clinical trial setting, where it can be deployed alongside quantitative motor speech analysis. LE can be derived by having either lay listeners or trained speech pathologists listen to speech recordings and attest to how much effort it took to understand the speaker (15, 18). To constrain some variability, our study employed clinicians trained in dysarthria and ALS to rate LE.

There were 125 participants in the LE Substudy (105 ALS and 20 controls), all of whom had at least two months of speech recording data, for a total of 2124 speech recordings (Table 2; Figure 1). Controls were matched on age and sex. Demographics for the LE Substudy participants are shown in Table 1. Each speech pathologist rated 2549 recordings (7647 total) including 20% that were presented twice to calculate intra-rater reliability. Of these recordings, 1953 recordings from 671 sessions passed quality control. Reasons for bad quality were wide-ranging and occurred at random.

**Table 2:** LE Speech Recording Sessions Collected, Quality Controlled, and Analyzed.

|  | **Patients** | **Controls** | **Total** |
| --- | --- | --- | --- |
| **Participants** | 105 | 20 (10 M, 10 F) | 125 |
| **Sessions before QC** | 584 | 124 | 708 |
| **Sessions after QC** | 552 | 119 | 671 |
| **Average Sessions per participant after QC** | 5.26 | 5.95 | 5.37 |
| **Timespan in study (days)** | 348 | 386 | 354 |
| **Unique Speech samples before Quality Control** | 1749 | 375 | 2124 |
| **Total Speech Samples Included after Quality Control** | 1605 | 348 | 1953 |
| **20% repetitions** | 325 | 78 | 403 |
| **Total Speech Samples** | 1930 | 426 | 2356 |

#### Intra- and Inter-Rater Reliability of LE is high

Intra-rater reliability, as measured by inter-class correlation (ICC), was excellent for two raters (0.92 and 0.91) and very good for one rater (0.89). The ICC did not differ significantly for each sentence’s task (11, 13, and 15 words). Pairwise inter-rater reliability ICC was excellent for two of the three rater pairs (0.92, 0.91) and very good for the third pair (0.88) (Figure 2).

**Figure 2.**
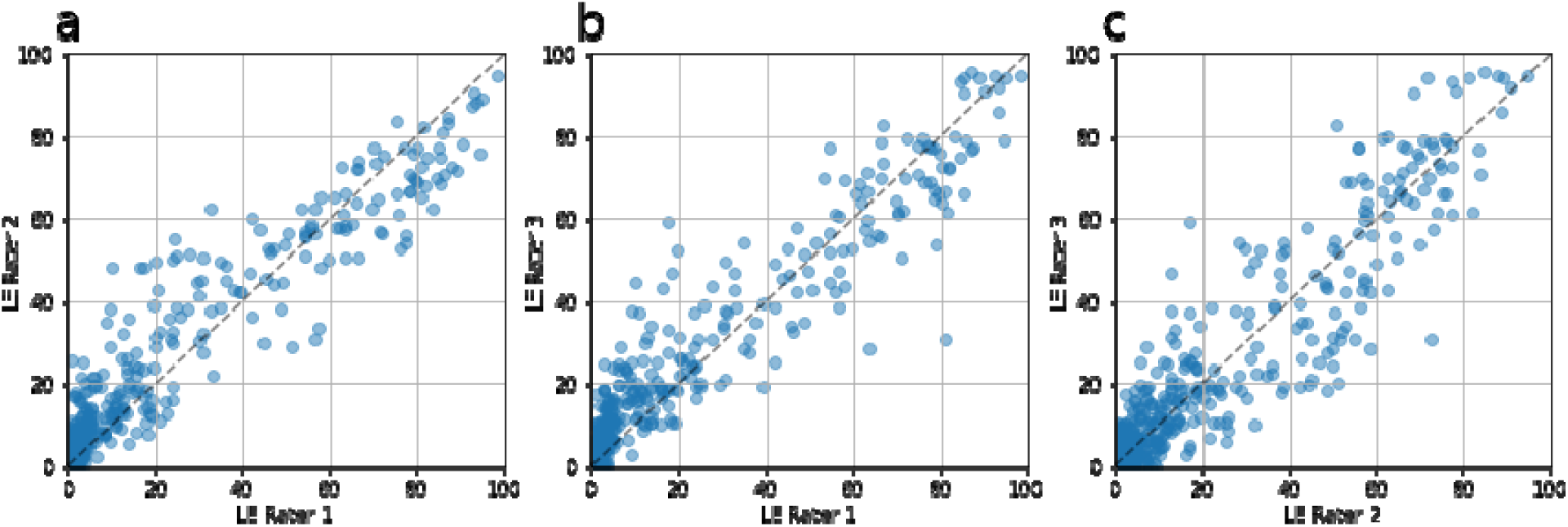
LE Inter-Rater Reliability. Pairwise inter-rater reliability was excellent for two of the three rater pairs: **(a)** LE Rater 1 and 2 ICC was 0.92; **(b)** LE Rater 1 and 3 ICC was 0.91. **(c)** Inter-rater reliability was very good for the third pair: LE Rater 2 and 3 ICC was 0.88.

#### LE and correlates with other features of bulbar function and declines as self-reported speech declines

There were moderate-high to high correlations between LE and other measures of ALS and dysarthria severity, suggesting that they measure related but not completely overlapping content. Mean LE correlated well with the Speech question (Q1) on the ALSFRS-RSE (Pearson R = - 0.72, p<0.001) and the bulbar subdomain (Q1-3) (R = −0.68, p<0.001). LE also correlated well with SR (R = −0.73, p<0.001) and AR (R=-0.75, p<0.001) (Figure 3a and 3b). SR and AR did not correlate as well with ALSFRS-RSE Q1 or the bulbar subdomain. (Figure 3a).

**Figure 3.**
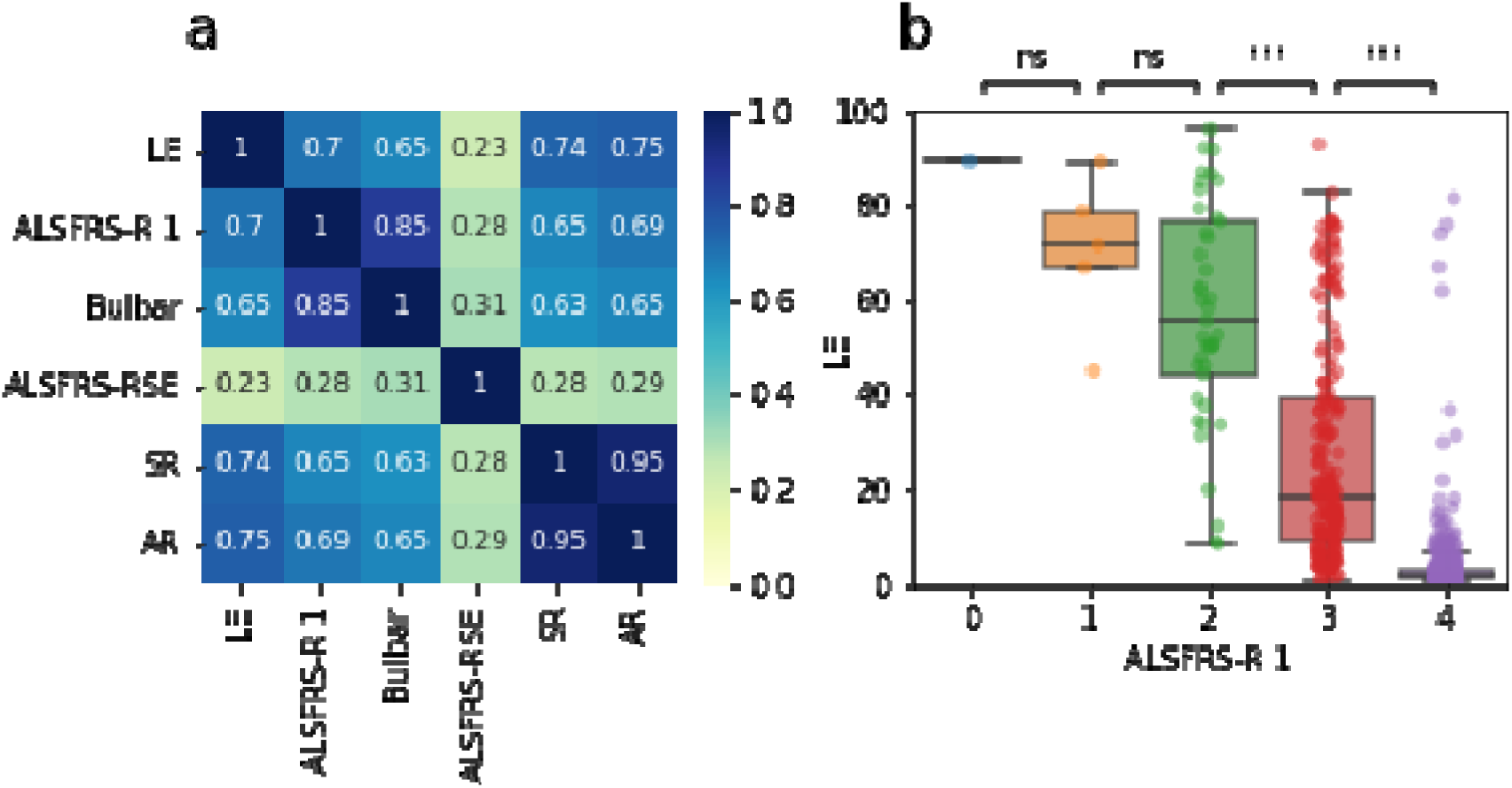
LE vs ALSFRS-RSE and acoustic features. **(a)** Correlation matrix was computed over the population of controls and PALS. Correlations are expressed in absolute values. Speaking Rate (words/min; SR) and Articulation Rate (syllables/sec; AR) are remarkably highly correlated (Pearson R = 0.95), indicating that they measure the same aspect of speech. There is a good correlation between Listener Effort (LE) and AR (Pearson R=-0.75), SR (−0.74), ALSFRS-R Question 1 (−0.70) and ALSFRS-RSE bulbar subdomain (−0.65). This indicates that while LE measures similar concepts to these measures, it also contributes non-overlapping information. As expected, because ALSFRS-RSE covers many more domains than just speech, it shows low to moderate correlation with AR, SR and LE. **(b)** In participants with lower self-reported speech function on the speech question (Q1) of the ALSFRS-RSE, LE increases (lower ALSFRS-RSE scores, and higher LE, denote lower speech function). Because Q1 of the ALSFRS-RSE only has five categorical answers, each category contains a wide spread of LE scores. These differences are significant comparing the categorical answers 2-3 and 3-4, but not 0-1 and 1-2. This may be because of the low numbers of participants in the lowest categories of ALSFRS-RSE Q1.

#### Unbiased clustering based on LE slope of decline defines distinct subgroups

We conducted an unbiased clustering of LE slopes using Mixture of Gaussian Processes (MoGP) on the 46 PALS with onset < 3 years prior to study entry and at least two audio sessions. The analysis revealed two distinct patterns of progression: non-progressors (cluster A, n=22, slope=0.05 pts/month, SD=0.40; cluster B, n=8, slope=0.03, SD=12.34), and progressors (cluster C, n=8, slope=3.26 pts/month, SD=9.18; cluster D, n=11, slope=3.76 pts/month; SD = 23.44) (Supplementary Figure 3). The unbiased identification of these clusters suggests that while the pace and pattern of dysarthria progression is variable, there are patterns to the progression.

#### PALS with bulbar involvement have similar rates of progression regardless of site of onset

The slope of decline of LE was 0.76 pts/month (SE=0.15, p<0.001) for PALS and 0.005 pts/month (SE=0.020, p=0.807) for controls (Figure 4a). PALS with bulbar onset ALS (n=23) had a faster LE progression rate (1.66 points/month) than non-bulbar onset (n=82) whose LE progression rate was 0.42 pts/month) (p<0.001; Figure 4b). Patterns were similar amongst PALS with onset <3 years prior to study entry, a population more resembling that of an ALS clinical trial population (Figure 4d and e).

**Figure 4.**
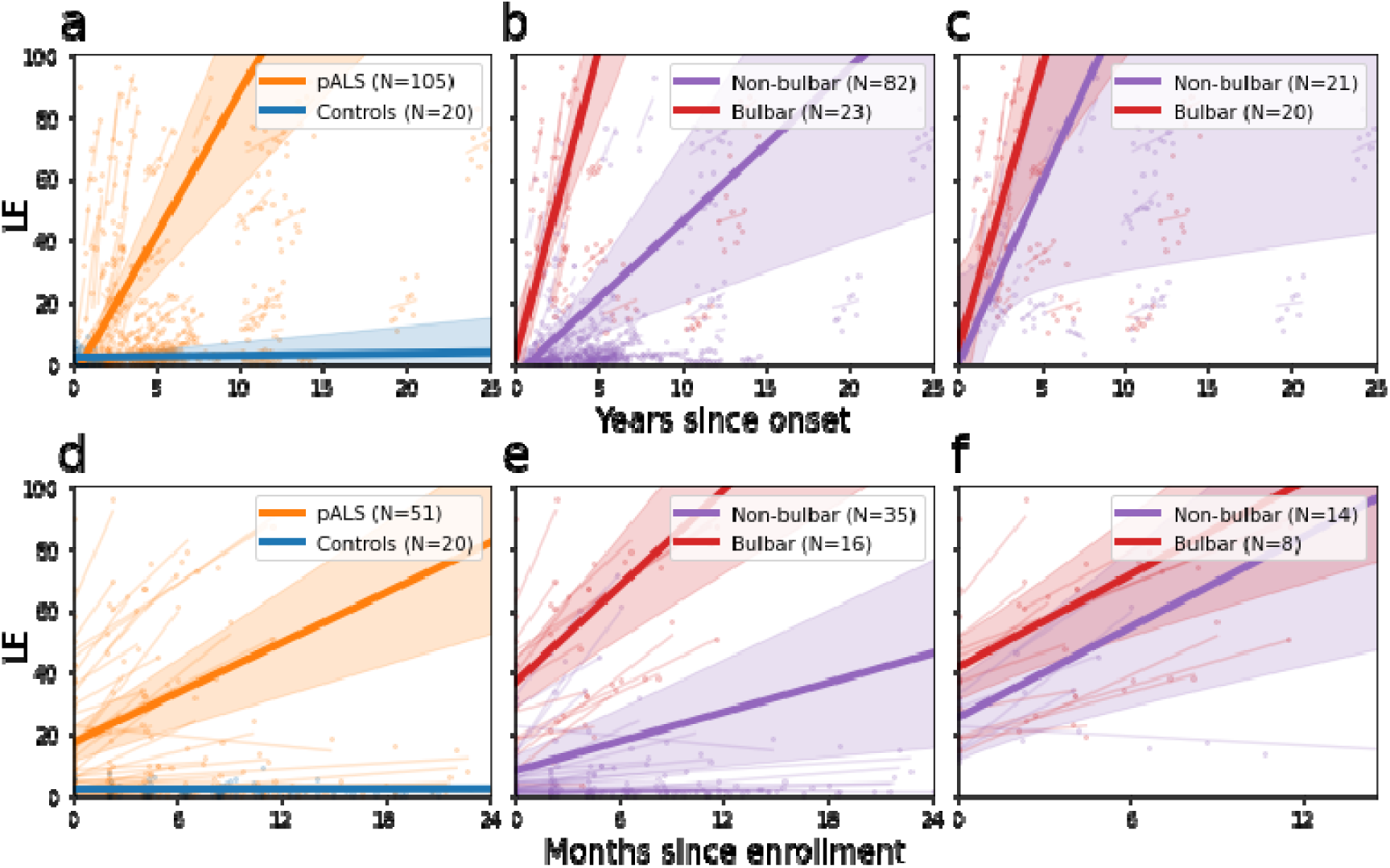
Progression of LE in PALS and controls. We analyzed ALS progression using linear mixed models (LMM) in different cohorts. In panels **(a-c)** we plotted LE data since onset from all participants, and in panels **(d-f)** we plotted LE data since enrollment from participants with onset of ALS within 3 years of study initiation. In panels **(a)** and **(d)** we compare PALS and controls. The slope of decline of LE is higher for PALS than controls in both all participants and those with onset <3yrs prior to enrollment. In panels **(b)** and **(e)** we compare PALS with bulbar and non-bulbar onset. PALS with bulbar onset show a faster slope of decline on LE than those with non-bulbar onset, consistent with the concept that PALS with bulbar onset have faster progression. In panels **(c)** and **(f)** we compare PALS with bulbar and non-bulbar onset, excluding PALS with LE scores in the normal range (0–10) at the time of enrollment. This partition focuses the analysis on PALS with current bulbar symptoms. In this analysis, LE slopes for PALS with bulbar and non-bulbar onset show no statistical differences. This suggests that once participants have developed bulbar symptoms, LE tends to progress at a similar rate, whether the disease began in the bulbar region or not.

#### Normal LE at baseline amongst PALS appeared to predict a group of participants likely to have slow progression

The LE slope for PALS with LE 0-10 at baseline (n=64) was only 0.03 pts/month, not significantly different than zero (p=0.13). When we included only PALS with onset within the last 3 years, mimicking a trial population, the progression rate for those with LE 0-10 at baseline (n=29) was 0.87 pts/month, which was significantly different than zero (p=0.027), but still slower than PALS with higher baseline LE (n=22) whose slope was 5.2 pts/month (p<0.001).

#### Regardless of site of onset, once bulbar symptoms begin, LE progresses at similar rates

We compared LE progression rates in PALS with bulbar onset and those with limb onset whose disease had progressed to involve bulbar function by the time of study enrollment. We dichotomized into PALS with bulbar onset (n=20) and those with limb onset. In PALS with limb onset, we included only those with abnormal LE scores (>10) at baseline in this study (n=21). The LE decline was not different regardless of site of onset (1.52 pts/month (bulbar onset; n=21), 0.98 pts/month (non-bulbar onset; n=20, p=0.36) This suggests that once participants have developed dysarthria, their LE scores tend to decline at a similar rate, whether ALS began in the bulbar region or limbs (Figure 4c and f). Again, the pattern was similar for PALS with symptom onset <3 years prior to study entry. As discussed in the prior section, a similar phenomenon was noted with SR.

#### LE is more sensitive to progression of bulbar symptoms over time than ALSFRS-RSE

In the LE substudy, 92 of the 105 PALS showed ALS progression (increase over time) in their LE score. By contrast, only 70 showed progression on Q1 of the ALSFRS-RSE, 64 on the bulbar subdomain, and 12 on SR, indicating that LE is more sensitive at detecting change over time than ALSFRS-RSE Q1, bulbar subdomain and SR (Table 3). We expected that the ALSFRS-RSE total score would identify the most progressors, since it measures progression in multiple functional domains. Accordingly, 95 PALS showed progression in ALSFRS-RSE total, which is a smaller increase over LE than we would have hypothesized.

**Table 3:** Comparison of the number of participants (n) with ‘Progression’ or ‘No Progression’ based on longitudinal changes in the ALSFRS-R Q1, Bulbar Subdomain, Total Score and Speaking Rate compared to LE.

|  |  | ALSFRS-R Q1 |  | ALSFRS-R Bulbar Subdomain |  | ALSFRS-R Total Score |  | Speaking Rate |  |
| --- | --- | --- | --- | --- | --- | --- | --- | --- | --- |
|  |  | NP | P | NP | P | NP | P | NP | P |
| LE | NP | 10 | 3 | 11 | 2 | 2 | 11 | 13 | 0 |
|  | P | 25 | 67 | 30 | 62 | 8 | 84 | 80 | 12 |

NP – No progression; P – Progression\*
\*Progression is defined as a score that declined more than the maximum change in the control population over the period of observation.

We compared the longitudinal coefficient of variation (CoV) of SR, ALSFRS-RSE Q1, and LE. Here, CoV is defined as (standard error of slope/mean of slope) for a variable. It allows comparison of the signal-to-noise of different outcome variables – CoV closer to 0 suggests less variability per unit change in the slope. It is one component of power calculations for planning clinical trials. Thus, outcome variables with lower CoV may provide higher statistical power in clinical trials. We evaluated CoV in the population of PALS who enrolled in the study within 3 years of symptom onset (a key enrollment criteria in many ALS trials; Figure 1). CoV was 0.40 for SR, 0.29 for ALSFRS-RSE Q1, and 0.20 for LE. For reference, the CoV for ALSFRS-RSE total score was 0.14.

### Model Prediction of LE

Given the strong performance of the speech-pathologist ratings of LE in evaluating dysarthria progression and its inherent clinical meaning, we built statistical models to predict LE Ratings. The model, called Listener Effort Prediction Model (LEPM), uses Lasso Regression on acoustical features.

An extensive list of acoustical features has been explored for the characterization of speech and speech disorders (27–29). Several promising candidate markers of bulbar motor decline were reported (12), from which we selected the following representatives of the phonatory, articulatory and resonatory speech subsystems: mean and standard deviation of pitch, mean and standard deviation of formants 1 and 2, standard deviation of the sound envelope, harmonic-to-noise ratio, shimmer, jitter and cepstral peak prominence (CPP) (30). We also included two representatives of the speech system: speaking rate and Whisper Confidence.

Whisper is a large-scale, weakly supervised speech recognition system trained on data sourced from the web (31, 32). The system’s primary function is to generate transcriptions from audio recordings containing speech. A modified version of this system also facilitates the extraction of timestamps corresponding to each transcribed word. Alongside each word, the system provides a confidence score reflecting the model’s certainty regarding the accuracy of the transcription for that specific word. By averaging the confidence scores of all words in a transcript, we derive a metric referred to as ‘whisper confidence.’ Finally, combining the number of words obtained in the transcript and the duration obtained from the timestamps, we computed the speaking rate.

#### LEPM predicts LE with high accuracy

We implemented a nested cross-validation scheme, with five outer-folds and five inner-folds within each outer fold. Each outer fold included 25 unique participants (PALS and controls). The model showed a robust performance: the root mean square error (RMSE), averaged across the five outer folds was 8.56 ± 0.60, and the average R² was 0.83 ± 0.07 (figure 5a). Notably, most of the predictive power of the LEPM comes from two key features: speaking rate and whisper confidence (figure 5b).

**Figure 5.**
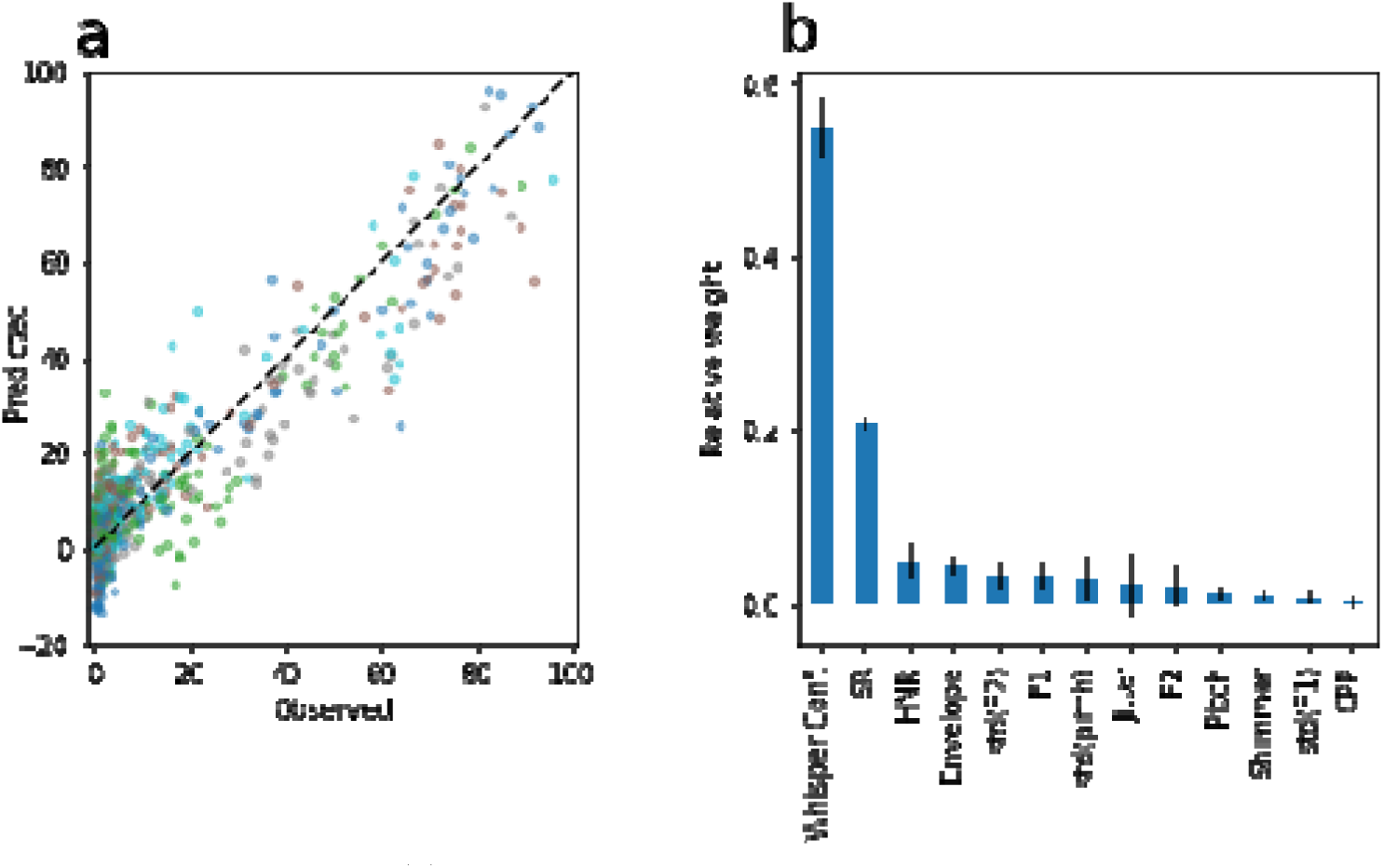
LE Prediction Model. **(a)** A Lasso regression model was trained and evaluated using nested cross-validation. The predicted LE output is plotted on the y-axis against the LE as assessed by SLPs on the x-axis. Each outer fold is shown in a different color. The model showed a robust performance: the RMSE averaged across the five outer folds was 8.56 ± 0.60, and the average R² was 0.83 ± 0.07. **(b)** The weight of each feature averaged across the outer folds (error bars are standard deviations). Most of the predictive power of the LEPM comes from Speaking Rate and Whisper Confidence

### The Everything ALS Speech Recording and Clinical Data Portal makes data and speech recordings available to researchers

All of the speech recordings and associated de-identified metadata are available to researchers via the Everything ALS Data Portal. The portal is hosted on Amazon Web Services in a HIPAA-compliant secure environment. Participants are identified in the study by a NeuroStamp, a coded identifier specific to the Everything Austen ALS Speech Study, that was derived from the NeuroGUID (global unique identifier). NeuroGUID is a coded identifier used to aggregate records from the same participants without the need for re-identification. NeuroStamp preserves the ability to aggregate data without exposing the NeuroGUID, adding an extra layer of de-identification.

Self-reported clinical data, including overall health information, ALS disease history, and routine outcome measures, including ALSFRS-RSE and speech recordings (uncompressed .wav format) are hosted in the data portal. Speech recordings and data can be requested via an online form at https://www.EverythingALS.org/available-data

## Discussion

This study has demonstrated that speech can be collected from a large cohort of participants with ALS over time using a remote study design with good participant retention and compliance, thanks to design and participant engagement elements of the study. Our results confirm prior findings that SR changes over time with ALS progression. However, SR captures only one aspect of the complex and variable dysarthria of ALS. Listener Effort (LE), a clinician reported COA, may provide a more holistic measure of effective communication in conditions with progressive and multifactorial impacts on speech. In this study, LE was more sensitive to change than the ALSFRS-RSE Q1 or the bulbar subdomain and other quantitative motor speech features. LE is practical to apply to ALS clinical trials as the assessment can be done on recordings by traditional cell phones, efficiently and reproducibly scored by SLPs using a simple online platform. While not tested in this study, we expect that LE could be implemented for global ALS trials, since SLPs who speak the same languages as trial participants are part of multidisciplinary care teams in other countries.

SLP ratings of LE showed very high intra- and inter-rater reliability. LE correlated with the bulbar subdomain of the ALSFRS-RSE and with other motor speech features, including speaking and articulation rates, but also added new information to the analysis. LE also provided interesting new insights into bulbar ALS progression. Most notably, when we excluded patients who had not yet developed bulbar symptoms at trial enrollment, the slope of LE decline was the same for participants who had bulbar or limb onset ALS. The same phenomenon was true for SR. This suggests that once bulbar symptoms start, *regardless of the initial site of ALS symptom onset*, dysarthria progresses at a similar rate.

In ALS clinical trials, where the goal of treatment is to slow or halt progression, outcome measures must record a decline over the period of observation. Those participants who do not show decline over the trial duration are non-informative and reduce trial power. Thus, measures more sensitive to change over time provide higher statistical power. LE detected change in bulbar function with more sensitivity than the ALSFRS-RSE bulbar subdomain, suggesting that it might add statistical power to clinical trial analyses relative to a bulbar subdomain analysis. Not only is LE a reliable, quantitative endpoint of bulbar function, it is also an inherently clinically meaningful endpoint. The effect of dysarthria on communication and, in turn, quality of life, is well accepted in clinical settings. The EFNS-ALS guidelines (33) suggest assessment of communication and treatment with communication support systems. People living with ALS worry about losing the ability to communicate (34, 35). Decreased speech function on the ALSFRS-R question 1 (speech), is associated with poorer quality of life (QoL) on the ALS-Specific QoL Questionnaire (ALSSQoL) (36). Furthermore, augmentative communication devices stabilize or improve both the quality of life and mood in people with dysarthria due to ALS (37, 38)Similar impacts of progressive dysarthria on quality of life have been demonstrated in Parkinson’s disease (39) and other neurological disorders (40).

Finally, our team generated a machine-learning model to predict LE, as rated by SLPs that was remarkably effective at predicting LE scores. Future studies to further validate the Listener Effort Prediction Model (LEPM) with external data sets are planned.

The Everything ALS Speech Data Repository (https://www.everythingals.org/available-data) is the largest ALS speech dataset available for broad use by ALS researchers and will facilitate method improvement and clinical trial modeling for the entire field. It will allow broader access to annotated speech recordings to hasten speech research, and benefit ALS trial design and drug development.

## Methods

The Everything ALS Speech Study was designed to gather speech recordings and patient reported outcomes (PROs) from people living with ALS to create a data and speech recording portal to share with the research community.

Enrollment in the study began in January 2021, and the study was designed to be fully remote to enable diverse and rapid enrollment. Participants were recruited through social media outreach, email outreach through ALS advocacy organizations, and the Everything ALS community webinars. Speech recordings included in this report were acquired between January 2021, and May 31^st^, 2023.

### Ethics Oversight

All aspects of the design and conduct of the Everything ALS Speech Study are done under the approval of the Western IRB. Every participant is presented with a digital written informed consent through a study portal and provides written documentation of informed consent prior to undergoing any study procedure.

All participants consented to inclusion of their study data (for example, coded identifiers, demographics, ALS history and outcome measures, and speech samples) in a large speech database that can be accessed by researchers under appropriate data use agreements.

### Compliance, Completeness and Study Dashboards

Non-interventional ALS studies consistently demonstrate approximately 50% loss-to-follow-up at six months (22, 41). Over the course of this study, the Everything ALS team developed participant retention strategies focused on platform improvements and frequent contact between the study team and participants. Participants received email reminders about their required sessions each week with the embedded secure link to record with one click. A programmable fit-for-purpose data dashboard was developed to allow 1) the Everything ALS study team to monitor enrollment, engagement, compliance, and data quality and 2) participants to track their own engagement and data. Participants could decide if they wanted to see their past ALSFRS-RSE score, with it displayed on a separate screen. Speech features from the reading passage, including speaking rate (wpm), diadochokinesis (DDK; “puh-tuh-kuh” (syllables per second)) and Loudness (dB, uncorrected for head-to-mic distance) were presented on an adjustable time graph to measure trend over time on the dashboard. Also, key statistics of the three features of last session, last 30-day average and all-time average were presented in a tabular form. The LE ratings were not presented to participants in this version of the dashboard, for practical reasons (LE was not rated in real time), to avoid the Hawthorne Effect, and because little was known about the clinical performance or meaning of the speech features at that time.

As a result of ongoing data quality and compliance review, university student volunteers were added as ambassadors to the Everything ALS study team to increase team outreach to participants, answer technical questions and encourage higher compliance. All student ambassadors undergo onboarding and compliance training, including for the remote delivery of the Edinburgh Cognitive Assessment Scale (ECAS) using a computer-based, examiner delivered form of the scale (42).

### Data Acquisition

The Everything ALS Speech Study uses a web-based platform from Modality.ai to present speech tasks to participants, an automated assistant to guide participants through the tasks, and video and audio to capture the results of speech tasks.

Speech and video recordings were obtained from participants as frequently as weekly with no restrictions on the number of recordings, although participants could record new sessions even if they missed prior sessions. ROADS were also collected through the platform until Oct 2022 where every other session was presented with the ROADS Self-entry form.

#### ALS Functional Rating Scale-Revised Self Entry

In this study, the self-entry form of the ALSFRS-R (ALSFRS-RSE) was presented to participants as a part of the Modality.ai data collection, along with speech recordings.

Recent data has demonstrated that the ALSFRS-R Self-Entry (ALSFRS-RSE) is very highly correlated with the ALSFRS-R at baseline (though ALSFRS-RSE is generally 1-3 points higher) and has similar slope of decline of the slope to the ALSFRS-R (4, 5, 43). The adoption of ALSFRS-RSE has allowed fully remote studies to follow the functional progression of people with ALS without the need for trained examiners, thus permitting more frequent sampling and thus smaller standard deviation on the slope of decline compared to traditional ALSFRS-R recording in clinic (44). Thus, while the ALSFRS-RSE score cannot be viewed as equivalent to the ALSFRS-R, it can be used as a proxy for the ALSFRS-R behavior. In this study, it is used to anchor speech analyses to functional status of participants.

The Rasch Overall Disability Scale (ROADS) (45) was delivered at alternating sessions until October 2022, when it was removed to reduce the burden of study visits. ROADS data are not presented in this manuscript but are available in the shared data.

### Data analysis

Demographics and compliance rates were tabulated using descriptive statistics.

We explored sensitive methods for detecting changes in speech. Speaking rate, a commonly used measure of dysarthria, is easily captured. We also explored several other measures of dysarthria, including diadochokinesis (DDK) the maximum rate that three syllables (i.e., “puh-tuh-kuh”) can be produced on a single breath. Quantitative motor speech analysis has become relatively common in ALS research, and this analysis generally followed previously published methods (4, 6, 8, 10, 11, 14).

Correlations between motor speech features and other clinical characteristics, such as ALSFRS-RSE were performed using Pearson R.

### Speech Recording Quality Assurance

Every session was screened for quality by the Modality.ai platform. Sessions were excluded if users experienced technical difficulties such as poor internet connection, session restarts or non-starts, device issues (eg. system errors), or premature session termination. Sessions were further screened for completeness. If a participant did not complete all the tasks in a session, the session is labelled incomplete. Additionally, sessions were discarded if they had either corrupted or non-existent WAV file.

In addition, all audio recordings were evaluated with Whisper for automated quality assurance. All audio recordings that matched at least 20% with the original text were included. This threshold was chosen because on direct listener evaluation, we found that, even in cases of severe dysarthria, Whisper recognized more than 20% of the original text. Thus, this threshold did not exclude severe dysarthria but did exclude some audio recordings that were technically inadequate for evaluation.

### LE Sub-study

As noted previously, the ALSFRS-R does not capture deep information about speech function, making it insensitive to the onset and progression of dysarthria. Examiner-rated outcomes of dysarthria severity could provide a sensitive and specific measure of progressive dysarthria.

In this sub-study, currently practicing speech and language pathologists (SLPs) with expertise in ALS, rated speech recordings from the Everything ALS Speech Study on a variety of outcome measures in a pilot study, from which LE was selected for formal study.

#### Pilot Studies

In the two pilot studies testing implementations of the SLP rating platform, three qualified, currently practicing speech and language pathologists with expertise in ALS care raters rated recordings from 10 participants performing three tasks: reading the bamboo passage (24), freely describing a line drawing picture (25), and reading three sentences (11, 13 and 15 words) (26). For each recording, each rater provided 11 outcome measures (LE, Overall Dysarthria Severity, Slow Speaking Rate, Voice Strain, Consistency, Reduced Intelligibility, Articulatory Imprecision, Dysphonia Severity, Hypernasality, Reduced Breath Support, Reduced Prosody).

#### Full LE Sub-study

Based on the pilots and bolstered by prior published experience, LE was selected as the outcome for the full sub-study to evaluate its performance as a quantitative measure of dysarthria progression in this population of people with ALS, and as a gold-standard benchmark against which to evaluate the performance of the quantitative motor speech features and complex predictive models.

The interface for performing LE was built using a web-based platform with secure login that presented raters with speech recordings one at a time (Supplementary Figure 4). First, a randomization system was designed to ensure recordings were presented in a random order from random participants. Second, at each session, a “warm-up” listening task was presented to the raters to ensure their equipment was working and they were ready for the tasks. Third, it was decided to repeat 20% of all recordings randomly across rating sessions to continue to assess intra-rater reliability. Fourth, a digital Visual Analogue Scale (VAS) was developed as a line with a slider that could be simply guided with a mouse or finger on a touch screen. The scale was 0 (easily understandable) to 100 (unintelligible even with full effort). Of note, during the pilot, the virtual visual analog slider used to select the LE score by each rater was initially positioned at 50 and did not require the rater to move the slider to record a score, which resulted in raters inadvertently scoring recordings a 50. For the full LE sub-study, the system was updated to require the slider to be moved prior to recording a score. One rater misunderstood, incorrectly assuming that the slider could not be moved back to exactly 50, thus, did not rate any recordings as exactly 50. Fifth, audio normalization was introduced. Sixth, all raters were provided with Sennheiser 280 Pro headphones. Seventh, all raters underwent hearing screening in the 60-70 dB SPL amplitude range. Finally, due to concerns about a potential ceiling effect in LE, a multi-talker babble noise (3 dB SNR) feature was built into the platform. When toggled on, this could introduce a layer of unintelligible babble to distract listeners. Because it was not found to impact intelligibility scores, it was not carried forward into the full sub-study.

For the full sub-study, participants with at least two months of speech recording data were included in the sample. The overall group of controls was selected to match PALS for mean age and sex. Three sentence recordings were selected for each session from each participant – sentences of 11, 13 and 15 words were used.

#### LE Analysis

Intra- and inter-rater reliability for LE was analyzed using interclass correlation (ICC). Intra-rater reliability was calculated using 20% of the audio recordings that were rated twice. Following existing guidelines, inter-rater reliability was calculated using all the audios (for those rated twice, we kept the first rating) (46). ICCs were interpreted as follows: < 0.5 poor reliability, 0.5 −0.75 moderate reliability, 0.75 - 0.9 good reliability, and > 0.90 excellent reliability.

The final LE for each participant in a session was computed as follows: Each session consists of three sentences. Three SLPs listened to each of these sentences, providing a total of nine ratings per session. The LE score for each participant’s session was determined by averaging these nine ratings.

Contingency tables were created to investigate the relative sensitivity to change of the LE metric, ALSFRS-RSE Q1, ALSFRS-RSE bulbar subdomain, ALSFRS-RSE total score, and Speaking Rate, contingency tables were created (Table 3). Change over time was dichotomized into either ‘Progression’ (the participant showed more decline than the maximum seen in the control population) or ‘No Progression’ (the participant did not show more decline than the max in the control population). The number of participants showing Progression was evaluated for each measure, with the measures identifying more participants with progression being the most sensitive to change.

Change in LE was evaluated using both unbiased and traditional modeling approaches. First, an unbiased machine learning method called Mixture of Gaussian Processes (MoGP) was used to identify groups of participants with similar progression rates on LE. MoGP identifies clusters of participants with similar rate and pattern of decline for a given outcome measure. It has been described for clustering participants based on ALSFRS-R decline (47).

To evaluate overall progression rate of LE, a Linear Mixed Model with ‘participant’ as Random Effect was applied. This model was used to determine whether the LE slope differs from zero. The study populations included a) controls, b) PALS, and c) PALS filtering those who had normal LE levels at the outset of the study. Further analysis within the PALS group was conducted using a Linear Mixed Model, again with ‘participant’ as Random Effect, to explore differences based on group assignment. Group comparisons included a) participants with bulbar-onset ALS versus non-bulbar-onset ALS to determine if the slopes of decline differ based on site of onset, and b) participants with bulbar-onset ALS versus non-bulbar-onset ALS excluding all participants with normal LE at the outset of the study, to determine whether, once bulbar symptoms have begun, the progression rate varies depending upon whether onset was bulbar or non-bulbar.

Longitudinal coefficient of variation (CoV) was calculated to enable a comparison of the residual error compared to the amount of change in outcome measures, including SR, ALSFRS-R Q1, and LE. CoV was also calculated for ALSFRS-RSE, as a point of reference. CoV could be calculated in several ways. Because the CoV in this case was being used to provide some insight into the performance of these variables as outcome measures in a clinical trial, CoV was calculated based on the linear mixed model slope estimate for each variable, such that CoV = Standard Error of slope/ Mean Slope. As such, the CoV allows comparison of the degree of residual variability standardized by the slope of change. Variables with CoV closer to 0 have less variability per unit change of the slope.

### LE Prediction Model

Again, given the robust performance of the LE, statistical models were built to predict LE. The model, called Listener Effort Prediction Model (LEPM), was built using a Lasso Regression model fed with acoustical features. Least Absolute Shrinkage and Selection Operator (Lasso) is a type of linear regression that incorporates a regularization through an L1 penalty (Regression Shrinkage and Selection via the Lasso, https://www.jstor.org/stable/2346178). This penalty term is proportional to the absolute value of the coefficients, leading to some coefficients being shrunk to zero. This model allows both predicting and identifying relevant features by excluding those with zero coefficients.

#### Acoustic Features

As noted, the set of standard features was chosen from a wide range of commonly reported markers in literature for tracking bulbar decline. These features include mean and standard deviation of pitch, mean and standard deviation of formants 1 and 2, standard deviation of the sound envelope, harmonic-to-noise ratio, shimmer, jitter and cepstral peak prominence (CPP). All these features were computed using the Parselmouth library (30). We also included two representatives of the speech system: speaking rate and Whisper confidence.

#### Model Evaluation

In our study, SLPs rated 708 sessions, from which we extracted features comprehensively. Following the quality assurance (QA) process, 671 sessions were retained for further analysis. A Lasso regression model was trained and evaluated using nested cross-validation with five outer-folds and five inner-folds within each outer fold. Each outer fold included 25 unique participants (PALS and controls). Uniformity of the target distribution across folds was confirmed via a Kolmogorov-Smirnov test between each outer fold and the remaining combined folds; this procedure was similarly applied within the inner folds, which comprised 20 unique participants each. Cross-validation within each inner fold determined the optimal level of regularization for the models. Subsequently, the best performing model from each outer fold was retrained using data from the other four outer folds and tested on the remaining fold.

## Data Availability

The repository of recorded speech and de-identified clinical data from this study is now available to ALS researchers to advance speech research in ALS (https://www.everythingals.org/available-data).

## Author Contributions (CRediT Classification)

INB– Conceptualization, Funding Acquisition, Investigation, Project Administration, Writing – Original Draft; RN - Conceptualization, Formal Analysis, Investigation, Methodology, Validation, Writing - Review and Editing; EGR - Conceptualization, Data Curation, Formal Analysis, Investigation, Methodology, Software, Writing – Original Draft; JP - Conceptualization, Data Curation, Formal Analysis, Investigation, Methodology, Software, Writing – Original Draft; MAT - Conceptualization, Data Curation, Formal Analysis, Investigation, Software, Methodology, Project Administration, Writing – Original Draft; CA- Data Curation, Formal Analysis, Investigation, Writing - Review and Editing; DES - Data Curation, Formal Analysis, Investigation, Methodology, Software, Writing - Review and Editing; FA - Data Curation, Formal Analysis, Investigation, Writing - Review and Editing; IE - Data Curation, Formal Analysis, Investigation, Writing - Review and Editing; AT - Data Curation, Formal Analysis, Investigation, Writing - Review and Editing; DH - Data Curation, Investigation, Writing - Review and Editing; AW - Data Curation, Investigation, Writing - Review and Editing; KS - Data Curation, Investigation, Writing - Review and Editing; SS - Data Curation, Investigation, Writing - Review and Editing; JRG - Conceptualization, Investigation, Methodology, Writing Review and Editing; LWO - Conceptualization, Investigation, Methodology, Project Administration, Writing – Original Draft; EF - Conceptualization, Formal Analysis, Investigation, Methodology, Project Administration, Software, Writing - Original Draft; JDB - Conceptualization, Investigation, Methodology, Project Administration, Writing - Original Draft.

## Acknowledgements

Jordan Green’s work was supported by National Institute on Deafness and Other Communication Disorders Grant K24 DC016312. We would like to thank all the participants in this study who generously dedicated their time and effort.

