## Supplemental Figures for "Listener effort quantifies clinically meaningful progression of dysarthria in people living with amyotrophic lateral sclerosis"

### Supplementary Figures

| | Geography (All N=291) |
|---|---|
| **N** | **State** |
| 30 | New Jersey |
| 22 | California |
| 17 | New York |
| 14 | Pennsylvania |
| 12 | Illinois |
| 10 | Massachusetts |
| 9 | Texas |
| 8 | Washington, Maryland |
| 7 | Michigan, North Carolina, Tennessee |
| 6 | Indiana, Missouri |
| 5 | Oregon |
| 4 | Colorado, Minnesota |
| 3 | Florida, Arkansas, Virginia |
| 2 | Georgia, West Virginia, South Carolina, Arizona, Ohio, Washington, D.C., Connecticut, Oklahoma |
| 1 | Alaska, Vermont, Kentucky, New Hampshire, Maine, Kansas, South Dakota, Alberta (Canada), Louisiana, Manitoba (Canada), Nevada, Alabama |
| 78 | No data |

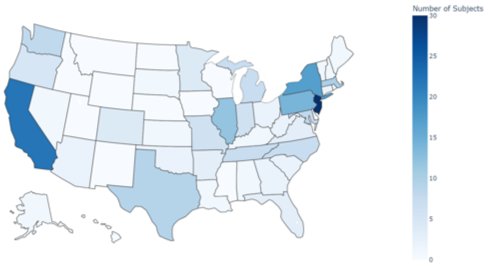


**Example**: there are three participants from Florida, three from Arkansas, and three from Virginia

### Supplementary Figure 1|Geographic distribution of the Speech Study

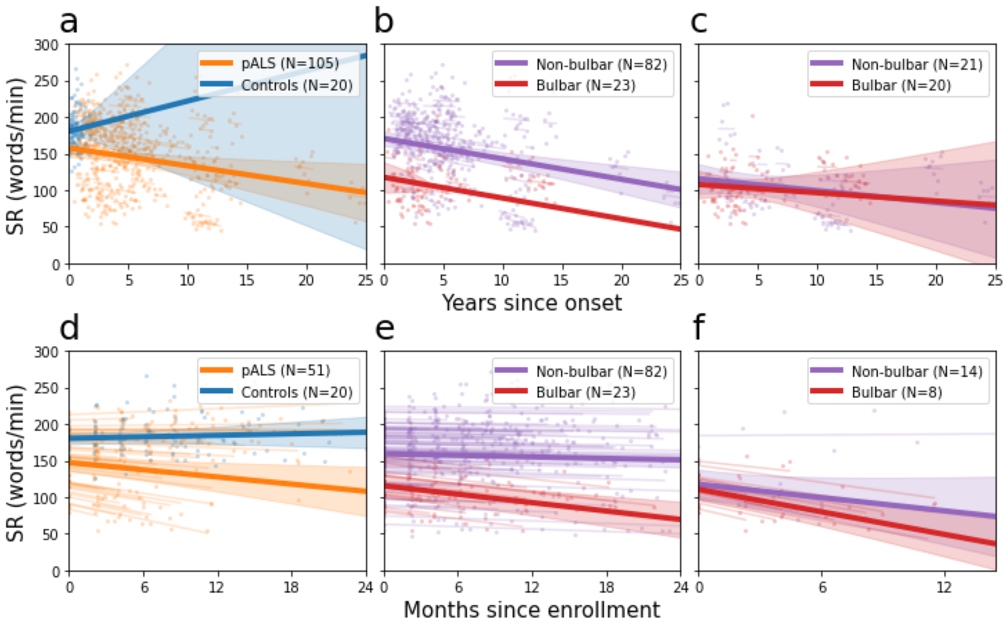


**Supplementary Figure 2 | Progression of Speaking Rate in PALS**. We analyzed ALS progression using linear mixed models (LMM) in different cohorts. In panels **(a-c)** we plot SR data since ALS onset from all participants, and in panels **(d-f)** we plot SR data since study enrollment for participants with onset of ALS within 3 years of study initiation. In panels (**a**) and **(d)** we compare PALS and controls. The slope of decline of SR is higher for PALS than controls in both all participants and those with onset <3yrs prior to enrollment. The positive slope and the dispersion of the regression for controls can be partly explained because the data from the controls are concentrated in a very short time range compared to the patients.. In panels **(b)** and **(e)** we compare PALS with bulbar and non-bulbar onset. **(b)** PALS with bulbar onset surprisingly show a similar slope of decline but a slight offset in y-intercept compared to those with non-bulbar onset when considering data since ALS onset, and **(e)** the expected faster slope of decline when considering only those with onset <3yrs prior to study enrollment. In panels **(c)** and **(f)** we compare PALS with bulbar and non-bulbar onset, excluding PALS with LE scores in the normal range (0-10) at the time of enrollment. This partition focuses the analysis on PALS with current bulbar symptoms. In this analysis, SR slopes for PALS with bulbar and non-bulbar onset show no statistical differences. In aggregate, this analysis still suggests that once participants have developed bulbar symptoms, SR tends to progress at a similar rate, whether the disease began in the bulbar region or not.

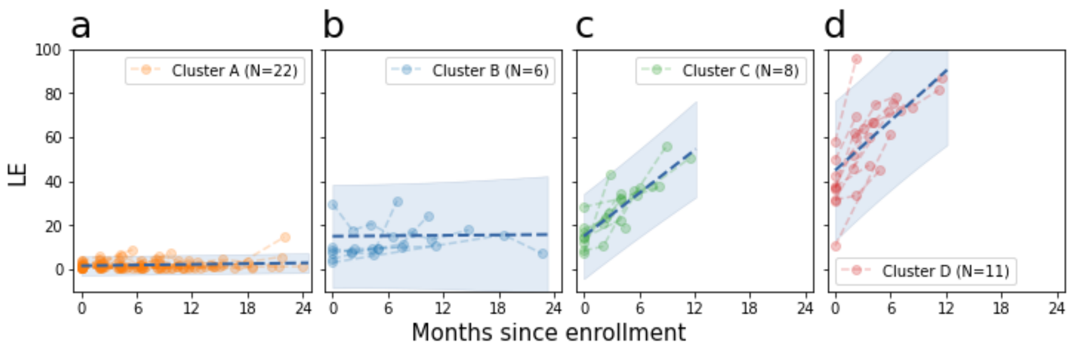


**Supplementary Figure 3 | Mixture of Gaussian Processes (MoGP) Clusters for LE**. Using the MoGP to cluster change in LE, four main clusters are created, roughly characterizing non-progressors **(a and b)**, those with slow change **(c)**, and those with faster change **(d)**. This suggests that even unbiased modeling approaches can identify clinically meaningful subpopulations within the larger group.

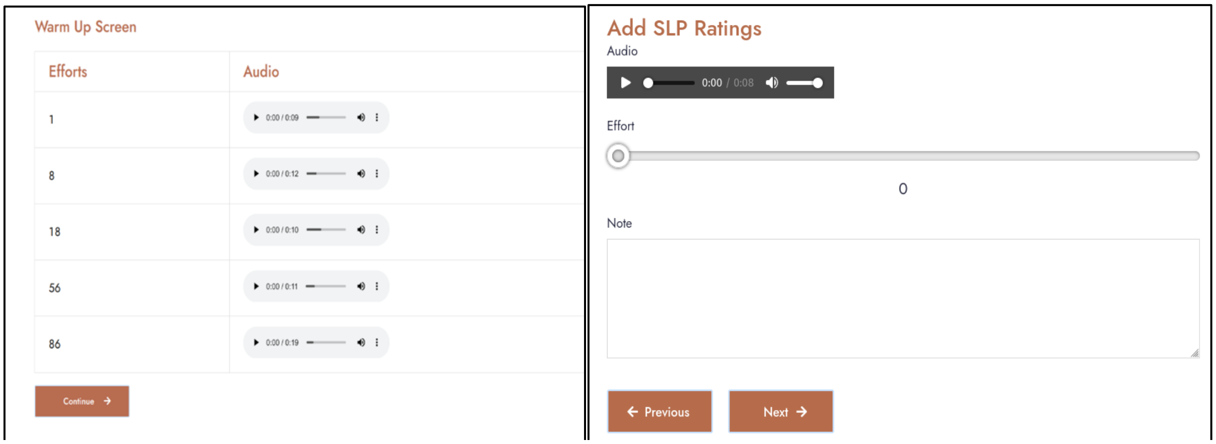


**Supplementary Figure 4 |** SLP Interface for Listening to Recordings and Rating LE
